# Enhancing Cause of Death Prediction: Development and Validation of ML Models Using Multimodal Data Across Multiple Healthcare Sites

**DOI:** 10.1101/2025.06.24.25330213

**Authors:** Mohammed Al-Garadi, Rishi J Desai, Kerry Ngan, Michele LeNoue-Newton, Ruth M. Reeves, Daniel Park, Jose J. Hernández-Muñoz, Shirley V. Wang, Judith C. Maro, Candace C. Fuller, Joshua Lin Kueiyu, Aida Kuzucan, Kevin Coughlin, Haritha Pillai, Melissa McPheeters, Jill Whitaker, Jessica A. Deere, Michael F. McLemore, Dax M. Westerman, Tony Morrow, Margaret A. Adgent, Michael E. Matheny

## Abstract

**Importance:** Timely and accurate determination of causes of death (CoD) is essential for public health surveillance, epidemiological research, and healthcare policy development. However, obtaining up-to-date and detailed CoD information is challenging due to delays in official death records and inconsistencies in data reporting across institutions.

**Objective:** To develop and validate machine learning (ML) models capable of predicting probable CoD by integrating comprehensive features from structured electronic health record (EHR) data, unstructured clinical notes, and publicly available data.

**Design, Setting, and Participants:** This multi-institutional retrospective cohort study was conducted at Vanderbilt University Medical Center (VUMC) and Massachusetts General Brigham (MGB). Deceased patients were included if they had at least one inpatient or outpatient encounter between October 1, 2015, and January 1, 2021, with corresponding death records from state health departments and the National Death Index. The study was comprised of 13,708 deceased patients from VUMC and 34,839 from MGB.

**Exposures:** Integration of structured EHR data, unstructured clinical notes processed using advanced language models, and publicly available data into machine learning models to predict CoD.

**Main Outcomes and Measures:** The primary outcome was the underlying CoD, classified into one of the top 15 National Center for Health Statistics (NCHS) rankable CoD categories, with all other causes grouped into an “Other” category. Model performance was evaluated using weighted area under the receiver operating characteristic curve (AUC) and weighted F-measure.

**Results:** The XGBoost model using structured EHR data alone achieved weighted AUCs of 0.86 (95% CI, 0.84–0.88) at VUMC and 0.80 (95% CI, 0.79-0.80) at MGB. Adding unstructured notes improved performance, with weighted AUCs of 0.90 (95% CI, 0.88–0.93) at VUMC and 0.92(95% CI, 0.91–0.92) at MGB. Adding publicly available data did not further improve performance. Cross-institutional validation revealed significant performance degradation.

**Conclusions and Relevance:** ML models integrating EHR structured and unstructured data to predict underlying CoD at the time of the most recent encounter among deceased patients achieved excellent performance within individual institutions. The inclusion of publicly available data did not improve performance, and all versions had poor portability between institutions. Healthcare institutions may benefit from adopting robust processes for locally tailored models, and future research should focus on enhancing model generalizability while addressing unique institutional data environments.

## Introduction

Timely and accurate identification of death and determination of causes of death (CoD) are vital for public health surveillance, epidemiological research, and healthcare policy development. Mortality is a critical outcome in medical product safety and effectiveness assessments, with all-cause mortality being one of the most studied outcomes in healthcare.^1–4^ Such mortality data inform strategies for disease prevention, resource allocation, and the evaluation of healthcare interventions. Obtaining up-to-date and detailed CoD information presents significant challenges due to delays in official death records and inconsistencies in data reporting across institutions. State-level mortality data can be delayed by 1 to 1.5 years, while national mortality data may lag by up to 2 years because of the time required for data collection, processing, and validation.^5,6^ This delay restricts its utility for analyses requiring more recent data ascertainment.^5,6^

Electronic health records (EHRs) offer a promising solution by providing a rich repository of patient information, including structured data—such as diagnoses, procedures, medications, laboratory results, and vital signs—and unstructured data like clinical notes. Leveraging EHR data for CoD prediction holds significant potential but presents challenges due to the heterogeneity of data sources and the complexity of integrating diverse data types.^7^

Previous studies to predict CoD using EHR data have often been limited by single-site data, smaller populations, or a focus on specific CoD, which may not generalize to broader, more diverse populations. ^8–13^ For instance, Kim et al.^9^ used a stacking ensemble method to predict CoD from patient checkup data, but their approach was limited by the exclusion of unstructured data and challenges in expanding to other institutional datasets.^9^ Jeblee et al. addressed CoD prediction using verbal autopsy (VA) narratives, applying natural language processing (NLP) techniques to unstructured narrative data. Their method utilized word frequency counts from free-text narratives to improve CoD classification accuracy, achieving 77% sensitivity for adult deaths across 15 categories. While their narrative-based approach outperformed structured data classifiers at the individual level, it was still limited to population-level estimations in low-resource settings.^8^ Additionally, the model’s reliance on the VA structure hindered its applicability to broader clinical datasets.

Our study represents a significant advancement by integrating both structured EHR data and unstructured clinical notes using advanced language models, resulting in a more comprehensive and scalable prediction system than those in previous studies. The inclusion of multiple multimodal data sources—including EHRs, clinical notes, and publicly available records—establishes a level of data integration not previously achieved, thereby extending the scope beyond prior research. Moreover, our study’s multi-institutional design encompasses a large and diverse patient population, thereby strengthening the analysis and conclusions drawn from extensive data.^8–13^

Integrating unstructured clinical narratives using advanced language model techniques can capture nuanced clinical information that is not readily available in structured data.^14–17^ Additionally, publicly available data sources, such as obituaries and memorial websites, may provide supplementary information that enhances mortality surveillance.^18–20^ Incorporating these varied data sources requires sophisticated feature engineering and validation against standardized reference-standard labels to ensure accuracy and reliability.

Recent advances in deep learning and NLP, especially transformer-based models like BERT and GPT, have improved the integration of unstructured data in healthcare.^21^ These models effectively process large volumes of clinical text, capturing complex linguistic patterns and medical terminology. By leveraging these advanced NLP and large language model (LLM) techniques, healthcare institutions can more readily incorporate unstructured data into predictive models, enhancing accuracy without extensive manual feature engineering.^22,23^ This scalability accelerates the development of predictive tools, making them more accessible for widespread clinical application and ultimately improving patient outcomes through informed decision-making.^23,24^

In this multi-institutional retrospective cohort study, we aimed to develop and validate machine learning models capable of predicting CoD by integrating comprehensive features from structured EHR data, unstructured clinical notes, and publicly available data. We hypothesized that integrating multimodal data sources with advanced NLP techniques would improve CoD prediction accuracy compared to models using structured data alone. Conducted at Vanderbilt University Medical Center (VUMC) and Massachusetts General Brigham (MGB), our study included a large patient population with a wide range of CoD. By utilizing standardized reference-standard death records from the National Death Index (NDI) and state health departments, we ensured reliable labels for model training and evaluation.

## Methods

### Study Design

This multi-institutional, retrospective cohort study aimed to develop and validate machine learning models for predicting CoD by integrating structured EHR data, unstructured EHR clinical notes, and publicly available data (Figure 1). Conducted at VUMC and MGB, the pipeline, as illustrated in Figure 1, incorporated feature engineering from structured EHR data and document-level embeddings from clinical notes using the Clinical Longformer model. Public data sources were categorized into top CoD groups and one-hot encoded with a large language model. These inputs were combined into machine learning models, trained using gold-standard death records from state health departments and the NDI, and cross-evaluated across both institutions for portability capacity and generalizability.

**Figure 1:**
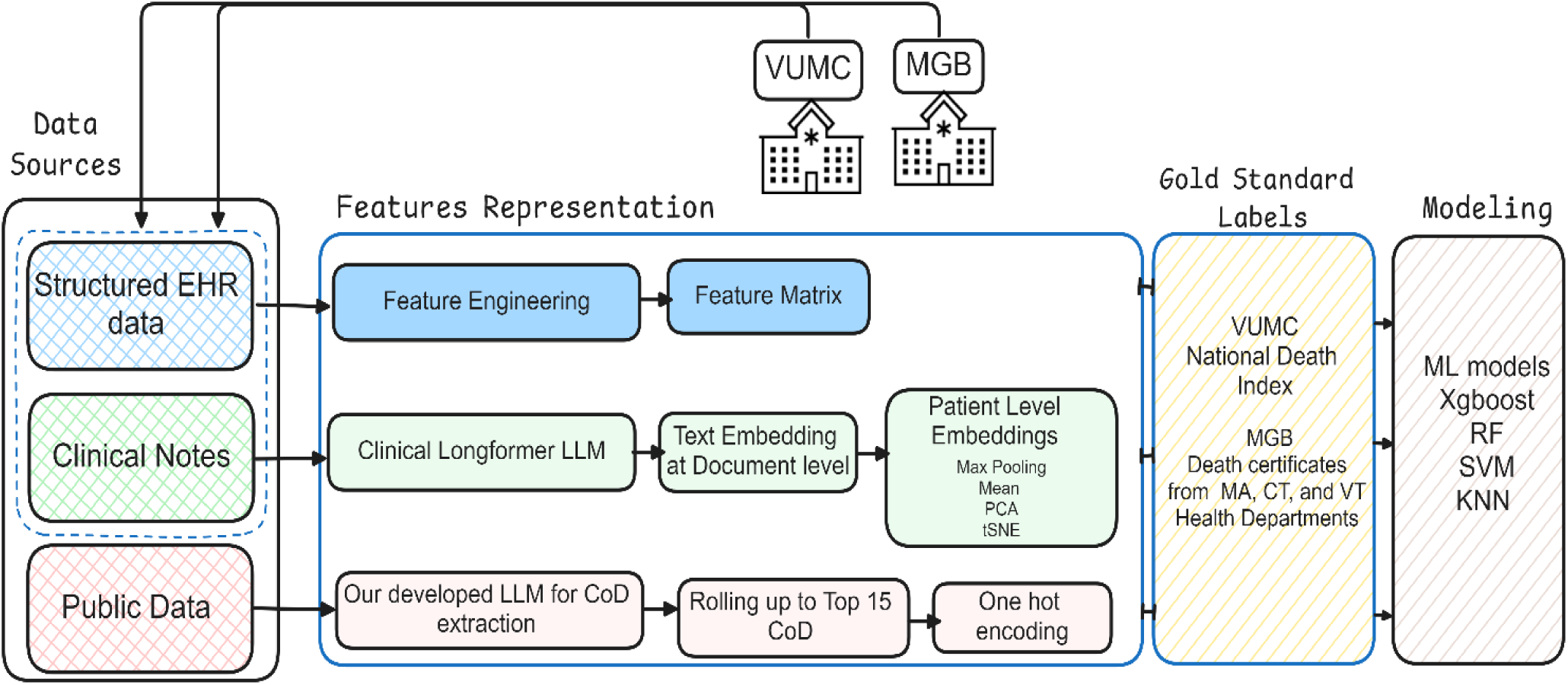
Pipeline for CoD Prediction Using Multimodal Data Sources and Machine Learning Models

### Study Population

We included retrospective cohorts from both MBG and VUMC in this study. For MGB, patients whose deaths were recorded between January 1, 2015, and Dec 31, 2020, in one of 3 state vital statistic database (Massachusetts, Connecticut, and Vermont) were eligible if they had ≥1 visit recorded in MGB EHRs within 180 days prior to death. Inclusion also required at least one clinical note within a six-month look-back period prior to the last healthcare system encounter. For VUMC, patients were included if they had at least one inpatient or outpatient encounter within the same date range, matched to the NDI with a recorded date of death between January 1, 2019, and December 31, 2021. They were also required to have at least one inpatient or outpatient encounter documented in the EHR within the six-month look-back period prior to their last recorded encounter.

### CoD Outcome Definitions

The outcomes of interest were the causes of death. These data are reported as part of the death certificate submissions in each state following the death of the patient. To align with how vital statistics data are commonly reported and used in observational research, we classified CoD into the 15 most frequent National Center for Health Statistics (NCHS) rankable categories, with all other causes grouped into an “Other” category.^25^

### Reference Standard Data Collection

The reference standard for CoD was the official death record from the NDI for VUMC and the state health departments of Massachusetts, Connecticut, and Vermont for MGB. These records provided definitive and reliable sources for determining the primary CoD.^26,27^ The term “reference -standard death records” refers specifically to this authoritative mortality data from the NDI and state health departments.

### Data Sources & Feature Engineering

We collected data elements from the EHRs of both VUMC and MGB. Table 1 summarizes the model’s integrated data layers: structured EHR data (ICD codes, labs, meds, vitals), unstructured EHR notes (language model embeddings), and public data (top causes of death from the National Center for Health Statistics (NCHS)^25^ groupers).

**Table 1.**
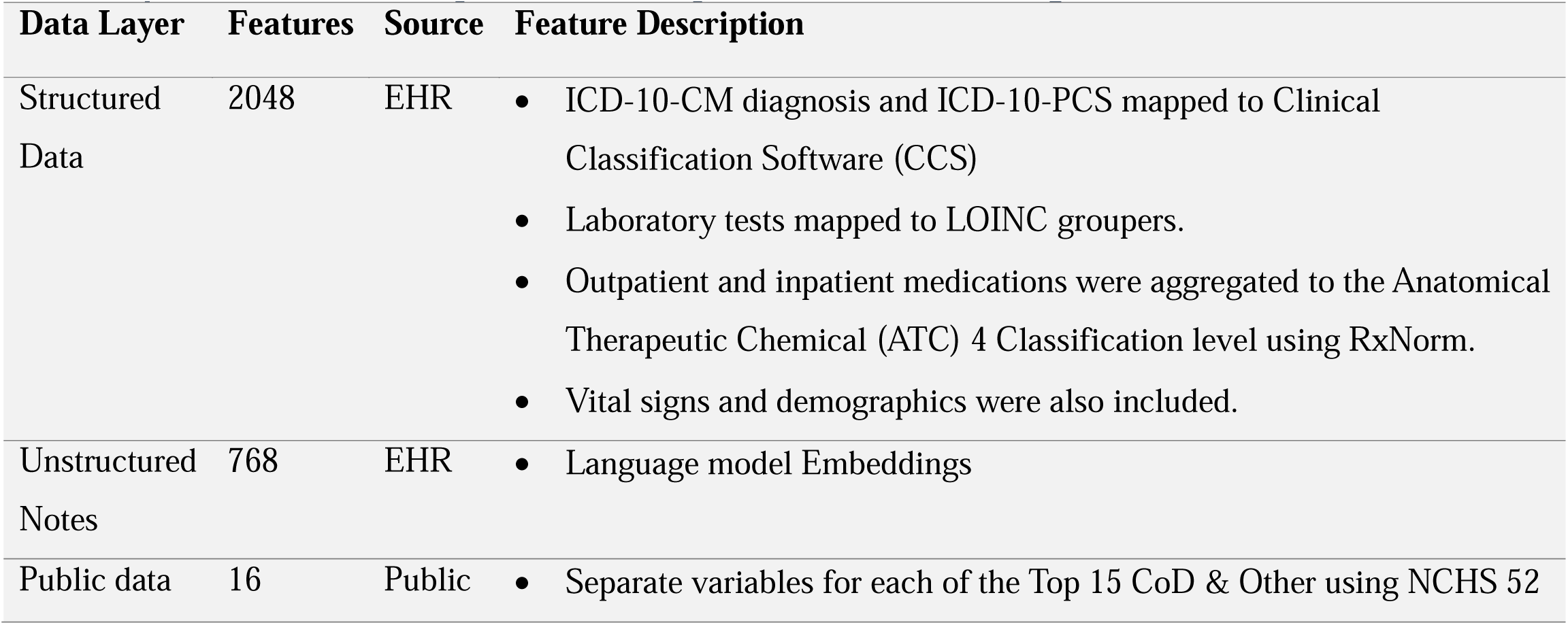
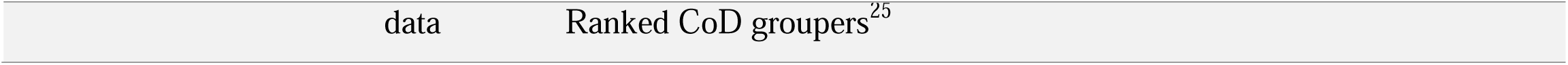
provides a summary of the data layers and features integrated into the model.

First, **structured data** elements were extracted that included diagnoses, procedures, medications, laboratory orders and results, vital signs, and demographic information. We transformed these sparse data into clinically meaningful groupings to reduce sparsity. Accordingly, we mapped ICD-10-CM diagnosis codes and ICD-10-PCS procedure codes to the Clinical Classification Software categories.^28^ Next, we grouped laboratory tests using the Regenstreif Logical Observation Identifiers Names and Codes [LOINC] test grouper mappings.^29^ We then transformed the medications into a RxNorm ingredient level binary yes/no exposure at the time of the last encounter and aggregated these data to the Anatomical Therapeutic Chemical [ATC] classification level 4.^30^ We also included the following vital signs: oxygen saturation, temperature, systolic and diastolic blood pressure, respiratory rate, and heart rate), and demographic data (Race, Gender, Ethnicity). This resulted in a total of 2,048 structured features.

Second, **unstructured data** comprising clinical notes were collected from the six months preceding the last patient encounter. We derived 768 unstructured features from embeddings of clinical notes using the Clinical Longformer small language model,^31^ which can process up to 4,096 tokens per document.

Third, we collected **publicly available data** from obituaries, crowdfunding platforms like GoFundMe, and memorial websites: Everloved and TributeArchives. A summary of the data characteristics of these data sources is noted in our previous paper.^32^ We derived 16 binary public data features representing the top 15 CoD categories defined by the NCHS^25^ and an “Other” category. CoD from public data was extracted and categorized using the LLaMA 3 LLM, leveraging our developed model to ensure alignment with standardized definitions.^32^

### Data Source Linkage

All data sources were linked using unique patient identifiers, resulting in an integrated dataset that included three types of features. The integration of public data with each institution’s EHR employed a method described in our previous paper.^32^ Public data, such as obituaries and memorial websites, were first downloaded and cleaned offline to ensure the data was processed in a controlled environment. The linkage between public data and EHR data was conducted on a local, secure server without exposure of the data to the internet, following strict data privacy protocols. We matched decedent information extracted from social media sources to patient records in the EHR using probabilistic linkage algorithms (SPLINK) based on first name, last name, date of birth, and state of residence. The linkage process demonstrated high accuracy, with exact matches on these variables indicating the reliable performance of the matching methodology.^33^ By linking and managing all data within each healthcare institution, we adhered to both institutional standards for patient privacy.

### Feature Selection

To enhance model interpretability and prevent overfitting, we applied feature selection techniques. Stability selection^34^ utilized base estimators like Lasso^35^ or Random Forest^36^ to identify features consistently associated with the outcome across subsamples. Random Forest feature selection involves extracting feature importance scores from trained Random Forest models. Additionally, we employed SelectFromModel using XGBoost’s feature importance metrics to retain the top features.^37^

### Model Development

At each site, ML models were developed to predict the top 15 underlying CoD categories and evaluated stepwise for three levels of feature inclusion (EHR structured data, EHR structured and unstructured data, and EHR structured and unstructured data with publicly available data). First, we assessed several machine learning algorithms—including XGBoost,^38^ Random Forest,^36^ Support Vector Machine (SVM),^39^ and K-Nearest Neighbors (KNN)^40^ methods in the structured EHR data. Performance comparisons were conducted to determine the optimal modeling method, which was subsequently applied to all remaining experiments across both study sites.

Second, to process unstructured EHR data, multiple ML methods were used for aggregating individual document-level embeddings into patient-last-encounter-level representations: max pooling, mean pooling, and dimensionality reduction techniques such as Principal Component Analysis (PCA)^41^ and t-distributed Stochastic Neighbor Embedding (t-SNE).^42^ Features were eventually added to the ML model that was determined to have the highest predictive performance, measured by the best F1-score and AUC-ROC, on the structured EHR data in the first step.

Thirdly, we evaluated whether the inclusion of cause of death information extracted from publicly available data using language models as described above would improve the performance of the selected ML modeling method using the pre-specified structured data feature management for both structured and unstructured EHR data.

### Statistical Analysis

Site specific models were initially assessed at both institutions. The dataset was split into 3 sets: 10% for hyperparameter tuning, 70% for training, and 20% for testing. Model performance was evaluated using the weighted area under the receiver operating characteristic curve (AUC) and the weighted F-measure. AUC^43^ incorporates class weights to address dataset imbalance, and the weighted F-measure balances precision and recall across all classes while considering class frequencies.^44,45^

All models were trained and evaluated using causes of death derived from official death records— specifically, the NDI for VUMC and state health department death records for MGB—as the gold-standard reference.

Correlation analyses compared CoD categories derived from public data with those from official death records. Spearman’s rank correlation coefficient and Kendall’s Tau assessed the strength and direction of associations between the two ranking sources.^46^ A p-value threshold of <0.05 was used to determine statistical significance.

We then conducted cross-institutional validation to assess model performance and generalizability across different healthcare institutions. Models trained on data from one institution (either VUMC or MGB) were tested on data from the other without any retraining or fine-tuning. This approach allowed us to evaluate how well the models generalized to new patient populations and clinical environments that differed from those of the training institution.

## Results

Patient demographics and CoD distributions are summarized in Table 2. The study included 13,708 deceased patients from VUMC and 34,839 from MGB. Malignant neoplasms were the leading CoD at both institutions, accounting for 4,155 deaths (30.3%) among VUMC patients and 10,961 deaths (31.5%) among MGB patients. Diseases of the heart were the second most common CoD, representing 2,192 deaths (16.0%) at VUMC and 6,445 deaths (18.5%) at MGB. COVID-19-related deaths comprised 1,044 cases (7.6%) at VUMC and 1,135 cases (3.3%) at MGB.

**Table 2.** Distribution of Top 15 CoD* at VUMC and MGB.

| **Top 15 CoD** | **VUMC** |  | **MGB** |  |
| --- | --- | --- | --- | --- |
| **Disease** | Count | % | Count | % |
| **Malignant Neoplasm** | 4155 | 30.3% | 10961 | 31.5% |
| **Diseases Of Heart** | 2192 | 16.0% | 6445 | 18.5% |
| **Covid19** | 1044 | 7.6% | 1135 | 3.3% |
| **Unintentional Injuries** | 1042 | 7.6% | 1270 | 3.6% |
| **Cerebrovascular Disease** | 612 | 4.5% | 1329 | 3.8% |
| **Chronic Liver Disease and Cirrhosis** | 364 | 2.7% | 324 | 0.9% |
| **Chronic Lower Respiratory Disease** | 353 | 2.6% | 1296 | 3.7% |
| **Diabetes Mellitus** | 306 | 2.2% | 769 | 2.2% |
| **Nephritis, Nephrotic Syndrome, And Nephrosis** | 194 | 1.4% | 852 | 2.4% |
| **Influenza And Pneumonia** | 188 | 1.4% | 693 | 2.0% |
| **Septicemia** | 157 | 1.1% | 525 | 1.5% |
| **Intentional Self Harm** | 153 | 1.1% | 118 | 0.3% |
| **Parkinson Disease** | 131 | 1.0% | 446 | 1.3% |
| **Essential Hypertension and Hypertensive Renal Disease** | 129 | 0.9% | 331 | 0.9% |
| **Alzheimer's Disease** | 115 | 0.8% | 703 | 2.0% |
| **All Other NCHS CoD** | 2573 | 18.8% | 7642 | 21.9% |
| **Total** | **13708** | **100%** | **34839** | **100%** |

\*CoD was defined using official death records from the NDI for VUMC and from state health departments (MA, CT, VT) for MGB.

### Initial Model and Approach Selection

At **VUMC**, the XGBoost model demonstrated superior performance, achieving a weighted AUC of 0.86 (95% CI, 0.82–0.88) (Table 3). Random Forest achieved a weighted AUC of 0.79 (95% CI, 0.75–0.83). The SVM and KNN models exhibited lower performance, with weighted AUCs of 0.73 (95% CI, 0.70–0.76) and 0.65 (95% CI, 0.60–0.70), respectively. At **MGB**, the Random Forest model achieved the highest weighted AUC of 0.82 (95% CI, 0.82–0.83), slightly outperforming XGBoost, which had a weighted AUC of 0.80 (95% CI, 0.79–0.80). The SVM and KNN models showed lower performance at MGB as well, with weighted AUCs of 0.75 (95% CI, 0.74–0.75) and 0.61 (95% CI, 0.61–0.62), respectively. <u>Based on the overall performance</u> <u>across both institutes</u>, XGBoost was selected for subsequent analyses due to its superior performance at VUMC and competitive performance at MGB.

**Table 3.** Performance of Machine Learning Models Only Using Structured features at VUMC and MGB.

| **Model** | **VUMC Weighted AUC (95% CI)** | **VUMC Weighted F-measure (95% CI)** | **MGB Weighted AUC (95% CI)** | **MGB Weighted F-measure (95% CI)** |
| --- | --- | --- | --- | --- |
| **XGBoost** | 0.86<br>[0.82 - 0.88] | 0.74<br>[0.71 - 0.76] | 0.8<br>[0.79 - 0.80] | 0.38<br>[0.37 - 0.39] |
| **Random Forest** | 0.79<br>[0.75 - 0.83] | 0.72<br>[0.69 - 0.75] | 0.82<br>[0.82 - 0.83] | 0.41<br>[0.40 - 0.42] |
| **SVM** | 0.73<br>[ 0.70 - 0.76] | 0.59<br>[0.56 - 0.62] | 0.75<br>[0.74 - 0.75] | 0.39<br>[0.38 - 0.40] |
| **KNN** | 0.65<br>[0.60 - 0.70] | 0.51<br>[0.46 - 0.56] | 0.61<br>[0.61 - 0.62] | 0.27<br>[0.26 - 0.28] |

To enhance model performance by incorporating information from unstructured clinical notes, various methods for aggregating data features extracted at the document level into patient-level representations were assessed using the XGBoost model at both VUMC and MGB. At **VUMC,** max pooling of embeddings yielded the best performance, increasing the weighted AUC to 0.90 (95% CI, 0.88–0.93) (Table 4). Other aggregation methods, including mean pooling, PCA, and t-SNE, also improved performance but to a lesser extent. At **MGB,** the mean pooling of embeddings resulted in the highest performance, with a weighted AUC of 0.92 (95% CI, 0.91–0.92). Max pooling also improved performance, achieving a weighted AUC of 0.88 (95% CI, 0.87–0.88). PCA and t-SNE methods showed lower performance at MGB, with weighted AUCs significantly lower than those achieved with pooling methods. <u>Based on these results,</u> max pooling was selected as the aggregation method at VUMC due to its superior performance, while mean pooling was selected at MGB.

**Table 4.** Performance of XGBoost Model including the features extracted using Unstructured Data Aggregation Methods at VUMC and MGB.

| **XGBoost Model** | **VUMC**<br>**Weighted**<br>**AUC**<br>**(95% CI)** | **VUMC**<br>**Weighted**<br>**F-measure**<br>**(95% CI)** | **MGB**<br>**Weighted**<br>**AUC (95%**<br>**CI)** | **MGB**<br>**Weighted F-**<br>**measure (95%**<br>**CI)** |
| --- | --- | --- | --- | --- |
| Structured and<br>Unstructured Data<br>(Max Pooling) | 0.90<br>[0.88-0.93] | 0.79<br>[0.77-0.81] | 0.87<br>[0.87 - 0.88] | 0.53<br>[0.52- 0.54] |
| Structured and<br>Unstructured Data<br>(Mean) | 0.89<br>[0.86-0.92] | 0.78<br>[0.79-0.77] | 0.92<br>[0.91 - 0.92] | 0.61<br>[0.60 - 0.62] |
| Structured and<br>Unstructured Data<br>(PCA) | 0.87<br>[0.85-0.92] | 0.75<br>[0.72-0.78] | 0.63<br>[0.62 -0.63] | 0.22<br>[ 0.21 - 0.23] |
| Structured and<br>Unstructured Data<br>(tSNE) | 0.90<br>[0.88-0.92] | 0.78<br>[0.76-0.79] | 0.61<br>[0.61-0.62] | 0.20<br>[ 0.19 - 0.21] |

### Model Performance: Structured Data Alone vs. Integrated Structured and Unstructured Data at Both Sites

At both VUMC and MGB, integrating structured and unstructured data statistically improved model performance, as shown in Table 5. At **VUMC**, the combined model achieved a weighted AUC of 0.90 (95% CI, 0.88–0.93) compared to 0.86 (95% CI, 0.84–0.87) with structured data alone (Table 5). Similarly, at **MGB**, the combined model reached a weighted AUC of 0.92 (95% CI, 0.91–0.92), outperforming the structured-only model with a weighted AUC of 0.80 (95% CI, 0.79–0.80).

**Table 5.** Model Performance with Structured Data Alone vs Combined Data at VUMC and MGB.

| **Sites** | **VUMC Trained and Tested** | **MGB Trained and Tested** |
| --- | --- | --- |

| *Metrics* | Weighted<br>AUC | Weighted<br>F-measure | Weighted<br>AUC | Weighted<br>F-measure |
| --- | --- | --- | --- | --- |
| *Models* | (95% CI) | (95% CI) | (95% CI) | (95% CI) |
| Structured data<br>alone | 0.86<br>[0.84 - 0.87] | 0.74<br>[0.71 - 0.76] | 0.8<br>[0.79 - 0.80] | 0.38<br>[0.37 - 0.39] |
| Structured and<br>Unstructured Data | 0.90<br>[0.88 - 0.93] | 0.79<br>[0.77 - 0.81] | 0.92<br>[0.91 - 0.92] | 0.61<br>[0.60 - 0.62] |

On an individual CoD level, as shown in Table 6, the addition of unstructured data led to variable improvements across different categories in both sites. **At VUMC,** the incorporation of unstructured data yielded statistically significant improvements (based on non-overlapping confidence intervals) for several CoD: chronic lower respiratory disease (AUC: 0.69 [0.68-0.70] to 0.78 [0.77-0.79]), intentional self-harm (0.73 [0.71-0.75] to 0.82 [0.79-0.85]), and cerebrovascular disease (0.67 [0.65-0.69] to 0.75 [0.73-0.77]). Additionally, several other CoDs also demonstrated improvements with non-overlapping CIs. Changes in diabetes mellitus (0.82 [0.80-0.84] to 0.80 [0.78-0.82]) and heart diseases (0.99 [0.97-0.99] to 0.98 [0.97-0.99]) were not statistically significant. **MGB** demonstrated more substantial improvements, particularly in chronic lower respiratory disease (0.75 [0.71-0.77] to 0.91 [0.90-0.94]), diabetes mellitus (0.75 [0.70-0.77] to 0.93 [0.92-0.95]), Parkinson disease (0.63 [0.57-0.67] to 0.95[0.92-0.97]), and influenza/pneumonia (0.63 [0.61-0.70] to 0.93 [0.91-0.95]). All improvements at MGB were statistically significant based on non-overlapping confidence intervals.

**Table 6.** AUC for Individual CoD at VUMC and MGB.

| **Disease** | **VUMC Trained and Tested** |  |  | **MGB Trained and Tested** |  |  |
| --- | --- | --- | --- | --- | --- | --- |
|  | *Counts* | *\*S*<br>(95% CI) | *\*\*S+U*<br>(95% CI) | *Counts* | *\*S*<br>(95% CI) | *\*\*S+U*<br>(95% CI) |
| **Overall Weighted AUC** | Total:<br>13708 | 0.86<br>[0.84-0.87] | 0.90<br>[0.88-0.93] | Total:<br>34839 | 0.80<br>[0.79-0.80] | 0.92<br>[0.91-0.92] |
| **Malignant Neoplasm** | 4155 | 0.94<br>[0.92-0.93] | 0.95<br>[0.94-0.96] | 10961 | 0.83<br>[0.82-0.84] | 0.90<br>[0.89-0.90] |
| **Diseases of heart** | 2192 | 0.99<br>[0.97-0.99] | 0.98<br>[0.97-0.99] | 6445 | 0.91<br>[0.90-0.92] | 0.99<br>[0.98-0.99] |
| **COVID19** | 1044 | 0.81<br>[0.79-0.83] | 0.86<br>[0.85-0.87] | 1135 | 0.74<br>[0.69-0.75] | 0.89 [0.88-0.91] |
| **Unintentional injuries** | 1042 | 0.82<br>[0.81-0.83] | 0.86<br>[0.85-0.87] | 1270 | 0.72<br>[0.71-0.75] | 0.93<br>[0.92-0.95] |
| **Cerebrovascular disease** | 612 | 0.67<br>[0.65-0.69] | 0.75<br>[0.73-0.77] | 1329 | 0.89<br>[0.84-0.92] | 0.98<br>[0.95-0.99] |
| **Chronic liver disease and cirrhosis** | 364 | 0.88<br>[0.86-0.90] | 0.89<br>[0.87-0.91] | 324 | 0.78<br>[0.76-0.81] | 0.92<br>[0.90-0.93] |
| **Chronic lower respiratory disease** | 353 | 0.69<br>[0.68-0.70] | 0.78<br>[0.77-0.79] | 1296 | 0.75<br>[0.71-0.77] | 0.92<br>[0.90-0.94] |
| **Diabetes Mellitus** | 306 | 0.82<br>[0.80-0.84] | 0.80<br>[0.78-0.82] | 769 | 0.75<br>[0.70-0.77] | 0.93<br>[0.92-0.95] |
| **Nephritis, nephrotic syndrome, and nephrosis** | 194 | 0.96<br>[0.95-0.97] | 0.97<br>[0.96-0.98] | 852 | 0.68<br>[0.63-0.71] | 0.95<br>[0.94-0.97] |
| **Influenza and pneumonia** | 188 | 0.82<br>[0.81-0.83] | 0.86<br>[0.84-0.88] | 693 | 0.63<br>[0.61-0.70] | 0.93<br>[0.91-0.95] |
| **Septicemia** | 157 | 0.92<br>[0.89-0.95] | 0.91<br>[0.89-0.95] | 525 | 0.68<br>[0.59-0.79] | 0.95<br>[0.89-0.99] |
| **Intentional Self Harm** | 153 | 0.73<br>[0.71-0.75] | 0.82<br>[0.79-0.85] | 118 | 0.88<br>[0.82-0.89] | 0.98<br>[0.97-0.98] |
| **Parkinson disease** | 131 | 0.89<br>[0.85-0.93] | 0.89<br>[0.85-0.93] | 446 | 0.63<br>[0.57-0.67] | 0.95<br>[0.92-0.97] |
| **Essential hypertension and hypertensive renal disease** | 129 | 0.67<br>[0.62-0.71] | 0.76<br>[0.73-0.79] | 331 | 0.81<br>[0.77-0.82] | 0.97<br>[0.96-0.98] |
| **Alzheimer's Disease** | 115 | 0.75<br>[0.73-0.77] | 0.79<br>[0.77-0.81] | 703 | 0.65<br>[0.64-0.66] | 0.85<br>[0.84-0.86] |
| **All Other NCHS CoD** | 2573 | 0.77<br>[0.74-0.80] | 0.85<br>[0.83-0.87] | 7642 | 0.73<br>[0.72-0.74] | 0.88<br>[0.88-0.89] |

\*S=Structured data \*\*S+U= Structured +Unstructured

### Public Data Integration

Integration of public data across two major health systems revealed variable utility in CoD prediction modeling. At VUMC, linkage with public sources—including obituaries and memorial websites—identified 6,285 patients, of whom 810 (12.9%) had discernible CoD information; 526 cases mapped to the top 15 NCHS rankable categories. Similarly, at MGB, 2,298 patients were linked to public data, with 451 (19.6%) having identifiable CoD information, though only 137 cases corresponded to the top 15 NCHS categories. The incorporation of sixteen public data-derived CoD features at VUMC did not improve overall weighted AUC, and MGB showed no improvement, with decreases in predictive accuracy.

Both institutions demonstrated strong rank correlations between public and healthcare datasets (VUMC: Spearman’s ρ=0.7785, Kendall’s τ=0.7113; MGB: Spearman’s ρ=0.6969, Kendall’s τ=0.5652; all p<0.05), suggesting alignment in CoD distributions despite limited utility for enhancing predictive models. Detailed analyses of ranking correlations and predictive performance following public data integration are provided in the supplementary materials.

### Cross-Site Model Evaluation

When models trained at one institution were tested on data from the other, significant performance degradation was observed. The XGBoost model using structured data trained on VUMC data achieved a weighted AUC of 0.86 when tested on VUMC data but decreased when tested on MGB structured data (Table 7). Similarly, the model trained on MGB data had a weighted AUC of 0.80 on MGB data but dropped when tested on VUMC data. Cross-site testing of VUMC-trained structured plus unstructured data models were also conducted and no substantial increase in cross-site performance was observed.

**Table 7.** Cross-Institutional Model Performance ^§^.

|  | VUMC Trained Model |  |  |  | MGB Trained Model |  |
| --- | --- | --- | --- | --- | --- | --- |
|  | Full Model Tested VUMC (*S) | Full Model Tested MGB (*S) | Tested VUMC (**S+U) | Tested MGB (**S+U) | Full Model Tested MGB (*S) | Full Model Tested VUMC (*S) |
| **Weighted AUC** | **0.86** | **0.49** | **0.9** | **0.51** | **0.8** | **0.55** |
| Malignant Neoplasm | 0.94 | 0.48 | 0.95 | 0.57 | 0.83 | 0.68 |
| Disease of heart | 0.99 | 0.47 | 0.86 | 0.48 | 0.91 | 0.51 |
| COVID19 | 0.81 | 0.55 | 0.86 | 0.47 | 0.74 | 0.41 |
| Unintentional injuries | 0.82 | 0.49 | 0.75 | 0.5 | 0.72 | 0.51 |
| Cerebrovascular disease | 0.67 | 0.35 | 0.89 | 0.4 | 0.89 | 0.53 |
| Chronic liver disease and cirrhosis | 0.88 | 0.53 | 0.78 | 0.51 | 0.78 | 0.44 |
| Chronic lower respiratory disease | 0.69 | 0.46 | 0.8 | 0.52 | 0.75 | 0.46 |
| Diabetes mellitus | 0.82 | 0.49 | 0.97 | 0.48 | 0.75 | 0.47 |
| Nephritis, nephrotic syndrome, and nephrosis | 0.96 | 0.54 | 0.86 | 0.5 | 0.68 | 0.55 |
| Influenza and pneumonia | 0.82 | 0.51 | 0.91 | 0.53 | 0.63 | 0.57 |
| Septicemia | 0.92 | 0.67 | 0.82 | 0.63 | 0.68 | 0.59 |
| Intentional self-harm | 0.73 | 0.43 | 0.89 | 0.48 | 0.88 | 0.41 |
| Parkinson disease | 0.89 | 0.5 | 0.76 | 0.53 | 0.63 | 0.63 |
| Essential hypertension and hypertensive renal disease | 0.67 | 0.6 | 0.79 | 0.44 | 0.81 | 0.51 |
| Alzheimer | 0.75 | 0.51 | 0.85 | 0.48 | 0.65 | 0.46 |
| All Other NCHS CoD | 0.77 | 0.51 | 0.98 | 0.47 | 0.73 | 0.52 |

<sup>§</sup>Confidence intervals are excluded as model degradation renders these calculations unwarranted. \*S=Structured data \*\*S+U= Structured + Unstructured

Feature selection methods—including Stability Selection, Random Forest Selection, and SelectFromModel—were applied to enhance model interpretability and potentially improve cross-site performance. By reducing dimensionality to potentially address overfitting to a local environment, these methods improved the performance somewhat for certain causes of death. However, they did not substantially mitigate the performance degradation observed during cross-site evaluations uniformly across all causes (Supplementary Table S2).

## Discussion

In this multi-institutional retrospective cohort study, we developed and conducted both internal and external validations of machine learning models to predict CoD by integrating structured EHR data, unstructured clinical notes, and publicly available data sources. Utilizing data from VUMC and MGB, we established a comprehensive and scalable pipeline for feature extraction and model training that can be adapted by other healthcare institutions. Our findings demonstrate that integrating unstructured clinical data with structured EHR data enhances predictive performance within individual institutions and offers a framework for improved mortality surveillance. In addition, our findings support the need to locally tune or train the modeling pipeline to preserve performance.

A key strength of our study is the development of a robust pipeline capable of processing large-scale data from diverse sources. By efficiently converting structured data—such as diagnoses, procedures, and laboratory results—and unstructured data—such as clinical notes—into machine learning–ready features, we enabled scalable modeling of patient information. The use of advanced NLP techniques, including strategies to aggregate information from the document level to the patient level, facilitated the integration of heterogeneous data types. Additionally, the utilization of standardized, high-quality gold-standard labels from official death records ensured the reliability and validity of our predictive models. Importantly, this pipeline was successfully deployed at both VUMC and MGB, allowing for local model training while maintaining methodological consistency. This approach is particularly beneficial for institutions with privacy concerns or varying data governance policies, as it permits site-specific adaptations without compromising data security.

This work underscores the importance of moving beyond structured data features and attempting to incorporate unstructured data into predictive models. While unstructured information from free-text notes takes more effort in pre-processing compared to structured features alone, which are mostly readily available, the performance gain in prediction models can be substantial, as demonstrated at both institutions in our study. These improvements were particularly pronounced in specific cause-of-death categories, emphasizing the value of unstructured data in capturing nuanced clinical insights not easily available in structured formats. Additionally, we observed variation in model performance across CoD categories. Common and well-documented conditions (e.g., cancer, heart disease) had higher AUCs, while others (e.g., Alzheimer’s, self-harm) showed lower and more variable performance. This underscores the need for condition-specific validation and tailored model interpretation. Integrating clinical narratives represents a crucial improvement, as they frequently contain essential details that may provide additional clinical context to bolster predictive model accuracy.

Despite the improvements observed within individual institutions, we encountered substantial performance degradation during cross-institutional validation. Models trained at VUMC performed inadequately when applied to MGB data, and vice versa. This decline underscores the significant impact of local data characteristics—such as differences in EHR systems, coding practices, and patient populations—on model performance.^47,48^ To mitigate potential overfitting to institutional data, we evaluated various feature selection methods. These approaches improved the performance somewhat for certain causes of death but the cross-site performance drop was still observed. This finding reflects broader debates in the literature, where localized models are often preferred for their tailored accuracy, whereas global models offer broader but less precise generalizability.^48,49^ Rather than attempting to directly transfer predictive models across institutions, which often leads to performance degradation due to these local variations, institutions may achieve better outcomes by adopting a shared, robust modeling process.^49^ Such an approach allows for the development of models tailored to specific data while maintaining methodological consistency. By disseminating a standardized process, as demonstrated in this study, institutions can build more accurate and context-specific models, which aligns with prior research advocating for adaptable, locally optimized models over a one-size-fits-all solution.^47,50,51^

The integration of publicly available data sources, such as obituaries and memorial websites, did not significantly improve overall model performance. This likely represents both a variability in how patients and their human networks publicly share information but also reflects a low rate of matching due to limitations in patient identifiers. These data sources have been thought to be valuable supplementary sources for timely mortality surveillance, particularly in situations where official death records are delayed or incomplete. However, while additional studies are needed to further evaluate the capacity of these data for this purpose, in this two-institution study we were unable to find significant improvements through incorporation of these data.

Our study contributes to the growing body of literature emphasizing the importance of integrating structured and unstructured data in predictive modeling. The scalable and adaptable pipeline we developed offers a framework that can be implemented by other medical institutions to develop high-performing models tailored to local patient populations. This is particularly relevant for CoD prediction, where timely and accurate information is crucial for public health planning and resource allocation.

### Limitations

This study has several limitations that should be considered when interpreting the findings. First, the heterogeneity of data between institutions—due to differences in EHR systems, coding practices, and patient demographics—may have contributed to the poor cross-site model performance. Models trained on data from one institution did not generalize well to another, indicating that local factors significantly influence model performance. This lack of generalizability underscores the need for site-specific model calibration to optimize predictive accuracy, which is critical for effective deployment across diverse healthcare environments.

Second, the retrospective nature of data collection may introduce biases related to data availability and documentation practices during the study period. Variations in how data were recorded, missing information and differences in clinical documentation could affect the consistency and reliability of the input features used for model training and validation.

In this study, the use of state death data for validation at MGB versus the NDI at VUMC is a limitation that may have impacted model comparability. This variation in death data sources introduces potential inconsistencies, which could influence the accuracy of CoD labeling and model generalizability across sites.

### Future Directions

To address the challenges of generalizability and enhance the applicability of predictive models across different healthcare settings, several strategies warrant exploration. Federated learning presents a promising approach by enabling models to be trained collaboratively across multiple institutions without sharing sensitive patient data.^52^ This method allows for broader learning from diverse datasets while maintaining patient privacy, potentially leading to more robust models that generalize better across institutions.

Transfer learning and domain adaptation techniques also offer potential benefits. By leveraging knowledge gained from diverse datasets and allowing models to adjust to local variations, these techniques can improve model performance in new settings.^53,54^

Advanced LLMs capable of handling both structured and unstructured data through few-shot learning represent another promising avenue. A unified LLM could streamline data integration and enhance generalizability by capturing complex insights across diverse data types, reducing the need for complex data-specific pipelines.^55^

Additionally, cross-institutional training and continuous model updates with real-time data could enable models to adapt to evolving healthcare environments. This dynamic updating could improve long-term performance and ensure that models remain relevant as clinical practices and patient populations change over time.^56^

## Conclusion

Our study demonstrates that integrating structured and unstructured EHR data enhances the predictive performance of machine learning models for CoD prediction within individual institutions. The development of a scalable and adaptable pipeline facilitates local model training and may improve mortality surveillance efforts. However, significant challenges remain in achieving cross-institutional generalizability, underscoring the need for site-specific tuning or alternative strategies to address local data heterogeneity.

## Funding statement

This project was supported by Task Order 75F40123F19010 under Master Agreement 75F40119D10037 from the US Food and Drug Administration (FDA). FDA coauthors reviewed the study protocol, statistical analysis plan, and the manuscript for scientific accuracy and clarity of presentation. Representatives of the FDA reviewed a draft of the manuscript for the presence of confidential information and accuracy regarding the statement of any FDA policy. The views expressed are those of the authors and not necessarily those of the US FDA.

## Data Availability Statement

The data used in this study are derived from electronic health records maintained by Vanderbilt University Medical Center (VUMC) and Massachusetts General Brigham (MGB), as well as mortality datasets provided by the National Death Index and the state health departments of Massachusetts, Connecticut, and Vermont. Access to these data is restricted due to institutional data use policies and patient privacy regulations.

## Code Availability

The code can be downloaded at the following link: https://github.com/CPHI-TVHS/Enhancing-Cause-of-Death-Prediction

## S1: Supplementary: Public Data Integration Results

**Table S 1.** Overall weighted AUC AND AUC for Individual CoD at VUMC (S=Structured data S+U+PD= Structured +Unstructured +Public data)

| Disease | VUMC Trained and Tested |  |  |  | MGB Trained and Tested |  |  |  |
| --- | --- | --- | --- | --- | --- | --- | --- | --- |
|  | Counts | S | S+U | S+U+PD | Counts | S | S+U | S+U+PD |
| Overall Weighted AUC |  | 0.86 | 0.90 | 0.90 |  | 0.80 | 0.92 | 0.72 |
| Malignant Neoplasm | 4155 | 0.94 | 0.95 | 0.95 | 10961 | 0.83 | 0.90 | 0.81 |
| Diseases of heart | 2192 | 0.99 | 0.98 | 0.98 | 6445 | 0.91 | 0.98 | 0.71 |
| COVID19 | 1044 | 0.81 | 0.86 | 0.86 | 1135 | 0.74 | 0.89 | 0.87 |
| Unintentional injuries | 1042 | 0.82 | 0.86 | 0.89 | 1270 | 0.72 | 0.93 | 0.68 |
| Cerebrovascular disease | 612 | 0.67 | 0.75 | 0.76 | 1329 | 0.89 | 0.97 | 0.70 |
| Chronic liver disease and cirrhosis | 364 | 0.88 | 0.89 | 0.89 | 324 | 0.78 | 0.91 | 0.83 |
| Chronic lower respiratory disease | 353 | 0.69 | 0.78 | 0.78 | 1296 | 0.75 | 0.91 | 0.73 |
| Diabetes Mellitus | 306 | 0.82 | 0.80 | 0.84 | 769 | 0.75 | 0.93 | 0.72 |
| Nephritis, nephrotic syndrome, and nephrosis | 194 | 0.96 | 0.97 | 0.92 | 852 | 0.68 | 0.95 | 0.74 |
| Influenza and pneumonia | 188 | 0.82 | 0.86 | 0.84 | 693 | 0.63 | 0.93 | 0.65 |
| Septicemia | 157 | 0.92 | 0.91 | 0.90 | 525 | 0.68 | 0.94 | 0.62 |
| Intentional Self Harm | 153 | 0.73 | 0.82 | 0.82 | 118 | 0.88 | 0.97 | 0.47 |
| Parkinson disease | 131 | 0.89 | 0.89 | 0.89 | 446 | 0.63 | 0.94 | 0.82 |
| Essential hypertension and hypertensive renal disease | 129 | 0.67 | 0.76 | 0.76 | 331 | 0.81 | 0.96 | 0.62 |
| Alzheimer's Disease | 115 | 0.75 | 0.79 | 0.79 | 703 | 0.65 | 0.85 | 0.77 |
| All Other NCHS CoD | 2573 | 0.77 | 0.85 | 0.86 | 7642 | 0.73 | 0.88 | 0.61 |

### Comparative Analysis of Public Data Versus NDI/State Data on Causes of Death

**Figure s1:**
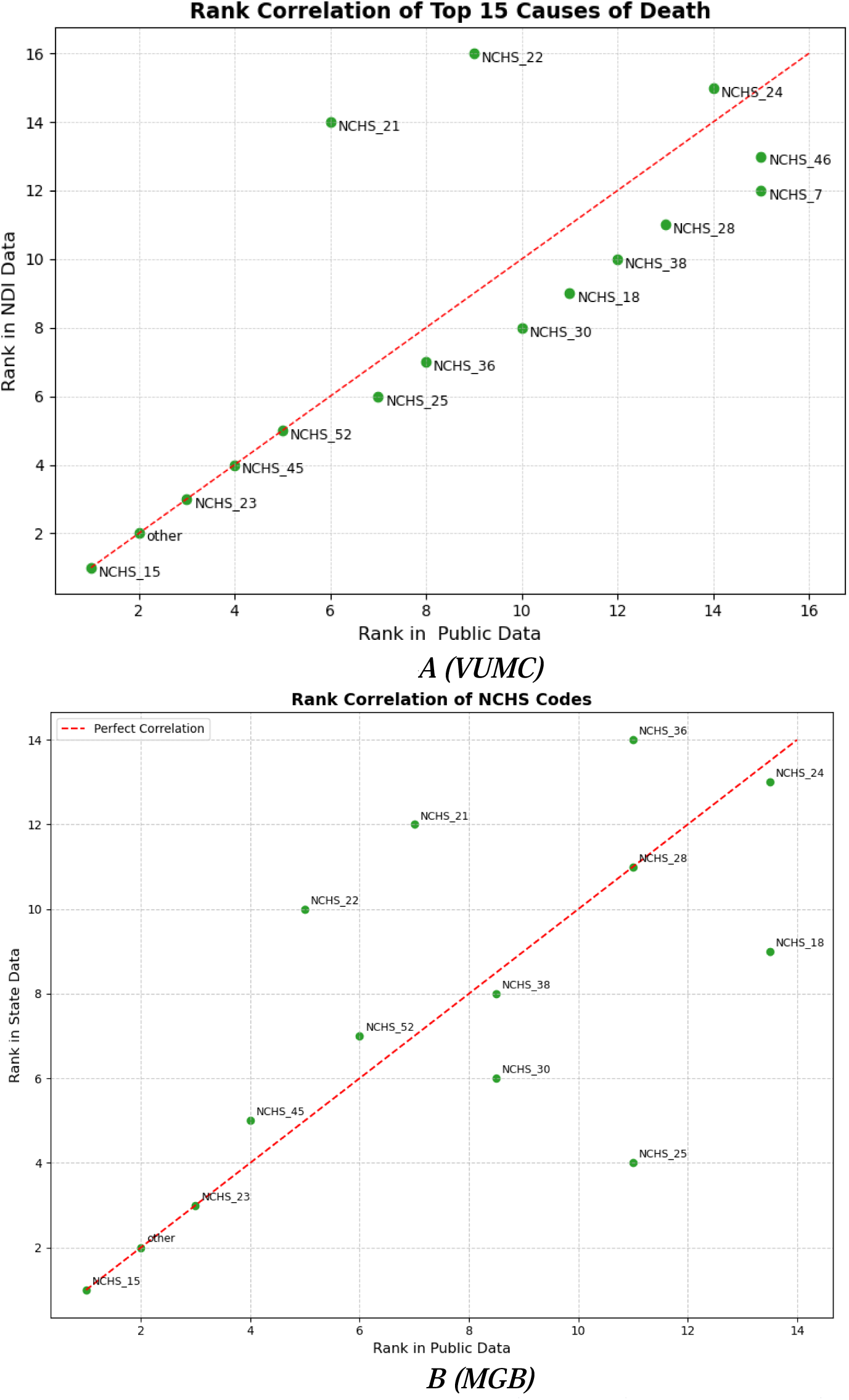
Rank Correlation of Top 15 Causes of Death - Public Data vs NDI Data. (NCHS_15 (Malignant Neoplasm), NCHS_23 (Diseases of Heart), NCHS_52 (COVID-19), NCHS_45 (Unintentional Injuries), NCHS_25 (Cerebrovascular Disease), NCHS_36 (Chronic Liver Disease and Cirrhosis), NCHS_30 (Chronic Lower Respiratory Disease), NCHS_18 (Diabetes Mellitus), NCHS_38 (Nephritis, Nephrotic Syndrome, and Nephrosis), NCHS_28 (Influenza and Pneumonia), NCHS_21 (Parkinson’s Disease), NCHS_24 (Essential Hypertension and Hypertensive Renal Disease), NCHS_22 (Alzheimer’s Disease), and Other (Other Causes)).

### Graph Overview

**Figure A (VUMC):** The scatter plot illustrates a strong positive correlation between the ranking of causes of death in public and healthcare data, as shown by points closely following a diagonal trend line. This alignment indicates that more frequently mentioned causes of death in public data, such as Alzheimer’s Disease and essential hypertension, also receive higher rankings in National Death Index (NDI) data. A few deviations from this trend, like Alzheimer’s Disease (NCHS_22) and essential hypertension (NCHS_24), show higher ranks in public data compared to healthcare records. **Figure B (MGB):** Similar to VUMC, the scatter plot for MGB shows a positive correlation between the ranks in public and healthcare data, suggesting alignment in cause of death distributions across data sources. However, there are minor deviations from the trend line, such as certain conditions ranking higher in state data than in social media mentions.

Together, Figures A and B suggest a strong rank correlation between public and institutional datasets across both VUMC and MGB, highlighting the consistency of cause of death distributions between public and healthcare records, with slight variations for specific conditions in each system.

### Spearman and Kendall Tau Correlations

**VUMC:** The correlation coefficients between public and healthcare data are high, with Spearman’s ρ = 0.7785 (p = 0.0004) and Kendall’s τ = 0.7113 (p = 0.0001), indicating statistically significant correlations. These values suggest a strong, consistent relationship between public reporting and healthcare data for CoD, implying alignment in cause-specific prevalence across sources.

**MGB:** Similarly, MGB demonstrates strong correlations, though slightly lower than VUMC, with Spearman’s ρ = 0.6969 (p = 0.0056) and Kendall’s τ = 0.5652 (p = 0.0058). These significant correlations reflect alignment between public perception and healthcare data at MGB, albeit with some variations in specific CoD rankings.

## S2: Supplementary: Feature selection results

Feature selection methods—including Stability Selection, Random Forest Selection, and SelectFromModel—were applied to enhance model interpretability and potentially improve cross-site performance. Despite reducing dimensionality and enhancing interpretability, these methods did not substantially mitigate the performance degradation observed during cross-site evaluations.

**Table S2.** Feature Selection Methods and Performance.

|  | VUMC Trained and Tested |  |  |  | VUMC Trained and MGB Tested |  |  |  |
| --- | --- | --- | --- | --- | --- | --- | --- | --- |
|  | Original Model | SFM | SS | RFS | Original | SFM | SS | RFS |
| **Weighted AUC** | **0.86** | **0.87** | **0.80** | **0.87** | **0.49** | **0.49** | **0.51** | **0.62** |
| Malignant Neoplasm | 0.94 | 0.95 | 0.92 | 0.94 | 0.48 | 0.53 | 0.55 | 0.65 |
| Disease of heart | 0.99 | 0.98 | 0.77 | 0.98 | 0.47 | 0.34 | 0.54 | 0.82 |
| COVID19 | 0.81 | 0.80 | 0.77 | 0.82 | 0.55 | 0.51 | 0.41 | 0.53 |
| Unintentional injuries | 0.82 | 0.81 | 0.77 | 0.82 | 0.49 | 0.55 | 0.47 | 0.48 |
| Cerebrovascular disease | 0.67 | 0.63 | 0.69 | 0.7 | 0.35 | 0.65 | 0.63 | 0.4 |
| Chronic liver disease and cirrhosis | 0.88 | 0.89 | 0.81 | 0.89 | 0.53 | 0.42 | 0.44 | 0.52 |
| Chronic lower respiratory disease | 0.69 | 0.68 | 0.65 | 0.66 | 0.46 | 0.52 | 0.54 | 0.43 |
| Diabetes mellitus | 0.82 | 0.82 | 0.75 | 0.79 | 0.49 | 0.53 | 0.46 | 0.51 |
| Nephritis, nephrotic syndrome, and nephrosis | 0.96 | 0.97 | 0.92 | 0.96 | 0.54 | 0.52 | 0.36 | 0.54 |
| Influenza and pneumonia | 0.82 | 0.78 | 0.75 | 0.76 | 0.51 | 0.55 | 0.38 | 0.58 |
| Septicemia | 0.92 | 0.91 | 0.84 | 0.92 | 0.67 | 0.55 | 0.49 | 0.66 |
| Intentional self-harm | 0.73 | 0.75 | 0.72 | 0.73 | 0.43 | 0.54 | 0.49 | 0.51 |
| Parkinson disease | 0.89 | 0.88 | 0.87 | 0.89 | 0.5 | 0.48 | 0.47 | 0.54 |
| Essential hypertension and hypertensive renal disease | 0.67 | 0.65 | 0.67 | 0.7 | 0.6 | 0.34 | 0.47 | 0.45 |
| Alzheimer | 0.75 | 0.74 | 0.70 | 0.75 | 0.51 | 0.52 | 0.46 | 0.51 |
| All Other NCHS CoD | 0.77 | 0.80 | 0.73 | 0.79 | 0.51 | 0.52 | 0.46 | 0.55 |

## Notes

**Funding sources:** The contents are those of the author(s) and do not necessarily represent the official views of, nor an endorsement, by FDA/HHS, or the U.S. Government. This project was supported by Task Order 75F40119F19002 under Master Agreement 75F40119D10037 from the US Food and Drug Administration (FDA).

### Competing Interest Statement

The authors have declared no competing interest.

### Funding Statement

Funding sources: The contents are those of the author(s) and do not necessarily represent the official views of, nor an endorsement, by FDA/HHS, or the U.S. Government. This project was supported by Task Order 75F40119F19002 under Master Agreement 75F40119D10037 from the US Food and Drug Administration (FDA).

### Author Declarations

The Institutional Review Board of Vanderbilt University Medical Center and the Institutional Review Board of Mass General Brigham gave ethical approval for this work.

### Summary of Updates

updated authors name and affiliation

